# Public health benefits of maternal influenza vaccination among pregnant women and infants <6 months in the United States, 2011–2020

**DOI:** 10.64898/2025.12.01.25339330

**Authors:** Sinead E. Morris, Alissa O’Halloran, Devi Sundaresan, Samantha M. Olson, Jessie R. Chung, Sascha Ellington, Catherine H. Bozio, A. Danielle Iuliano, Matthew Biggerstaff, Carrie Reed

## Abstract

**Background:** Pregnant women and infants <6 months are at increased risk of influenza-associated complications. Maternal influenza vaccination during pregnancy has a strong safety record and is effective in reducing the risk of illness in these groups. We estimate influenza disease burden, and the burden prevented by maternal influenza vaccination, among pregnant women and their infants <6 months in the United States during the 2011/12–2019/20 influenza seasons.

**Methods and findings:** We applied a multiplier model and compartmental framework to data that included monthly influenza-associated hospitalizations among pregnant women and infants <6 months, influenza vaccination coverage among pregnant women, and influenza vaccine effectiveness. We estimated the number of influenza-associated hospitalizations and intensive care unit (ICU) admissions among pregnant women and infants <6 months, and the corresponding number prevented by vaccination. We also simulated alternative vaccination coverage scenarios to investigate whether increased and/or earlier influenza vaccination uptake each season could have prevented additional hospitalizations. From 2011/12–2019/20, we estimated that 2,670–10,000 influenza-associated hospitalizations and 76–486 ICU admissions occurred annually among pregnant women, and 3,960–13,100 hospitalizations and 457–1,830 ICU admissions occurred annually among infants <6 months. Influenza vaccination prevented an estimated 33–2,440 hospitalizations and 1–166 ICU admissions annually among pregnant women, and 29–1,350 hospitalizations and 4–178 ICU admissions annually among infants <6 months. We projected that increased influenza vaccination uptake would have prevented more hospitalizations among pregnant women than earlier vaccination, whereas the converse was true for infants <6 months. Together, we estimated increased and earlier vaccination uptake could have prevented an additional 12–942 influenza-associated hospitalizations annually among pregnant women and 17–1,010 hospitalizations annually among infants <6 months.

**Conclusions:** Our findings suggest maternal influenza vaccination reduces disease burden among pregnant women and their infants <6 months who are too young to be vaccinated.

## Introduction

Pregnant women and infants <6 months are at increased risk of influenza-associated complications^1–6^. In the United States, the Advisory Committee on Immunization Practices (ACIP) and the American College of Obstetricians and Gynecologists (ACOG) recommend seasonal influenza vaccination for all women who are or might be pregnant during the influenza season to reduce the risk of respiratory disease^7–9^.

While the primary goal of influenza vaccination during pregnancy is to protect the health of pregnant women, protection can also be conferred to their infants through the transplacental transfer of maternal antibodies during fetal development^10,11^. Since infants are not eligible to receive influenza vaccination until 6 months of age, maternal immunization can provide protection during the early months of life when they are too young to be vaccinated^7,10,11^.

Influenza vaccination coverage among pregnant women ≥18 years increased from 47% in 2011 to 61% in 2020, but remained below the Healthy People 2030 Target of 70%^12,13^. Potential barriers to maternal vaccination include safety and financial concerns^14^. In addition, the opportunity for vaccination is likely influenced by the timing of pregnancy in relation to the timing of the influenza vaccination campaign^15^. The latter typically begins in September each year, although since 2021, ACIP has stated that vaccination can be considered as early as July or August for pregnant women in their third trimester to reduce the risks of influenza illness in their infants following birth^7^. Though the timing of the vaccination campaign is well-aligned to protect pregnant women from influenza during the season, infants who are born before the beginning of the vaccination campaign will miss an opportunity to be protected by maternal vaccination but may still be at risk of influenza during the upcoming season (before they are eligible for vaccination)^16^. Understanding the different impacts of influenza vaccination in preventing influenza-associated illness among pregnant women and their infants <6 months could help inform strategies to increase maternal influenza vaccination uptake and protection in these populations.

Here, we extend an established compartmental framework to estimate influenza disease burden, and the burden prevented by influenza vaccination, among pregnant women and their infants <6 months in the United States during the 2011/12 to 2019/20 influenza seasons^17,18^. We also investigate whether increased and/or earlier influenza vaccination uptake among pregnant women each season could prevent further disease burden among both groups. Our framework can help quantify the severity of seasonal influenza in pregnant women and infants <6 months, and the benefits of maternal influenza vaccination.

## Materials and Methods

### Surveillance data and model inputs

We collated demographic information to estimate the total number of pregnant women and infants <6 months in the United States from 2011–2020 (sources listed in the Supplementary Information). For infants, this included monthly numbers of live births and annual mortality rates for infants <1 year per 1,000 live births^19,20^. The latter were multiplied by one half to estimate annual mortality rates among infants <6 months (assuming equal mortality rates among infants <6 months and ≥6 months). The total number of infants <6 months was estimated as the total number of live births reported in the current and preceding 5 months, adjusted for infant mortality. For pregnant women, we obtained annual pregnancy rates per 1,000 females 15–44 years, the percentage of pregnancies ending in live birth, abortion, or fetal loss, and annual estimates of the total US population of females 15–44 years^20,21^. We then estimated the total number of pregnant women in each season by multiplying the annual US population of females 15–44 years by the annual pregnancy rates, adjusted for the relative frequency and duration of pregnancies ending in live birth, abortion, and fetal loss^22^.

For influenza surveillance data, we obtained monthly numbers of laboratory-confirmed influenza-associated hospitalizations and intensive care unit (ICU) admissions among pregnant women 15–44 years and infants <6 months for the 2011/12–2019/20 influenza seasons from the Influenza Hospitalization Surveillance Network (FluSurv-NET; Figure S1)^23–25^. FluSurv-NET conducts population-based surveillance each year from October to the following April and covers approximately 29 million people (or 9% of the US population). To account for under-ascertainment of laboratory-confirmed influenza-associated hospitalizations, we calculated season-specific under-detection multipliers that adjusted for FluSurv-NET testing frequencies and diagnostic test sensitivity^26^. This information was not directly available for pregnant women or infants <6 months and so we instead used information for women 18–49 years and infants <1 year, respectively. We also obtained estimates of the total FluSurv-NET catchment population of women 15–44 years and infants <1 year for each season. We translated the former to estimates of pregnant women in the catchment area by multiplying by the adjusted annual pregnancy rates, and the latter to estimates of infants <6 months in the catchment area by multiplying by one half (assuming equal representation of infants <6 months and ≥6 months).

In addition to hospitalizations and ICU admissions, we collated available information on influenza-associated symptomatic and medically attended illnesses. There were no direct sources of influenza-associated symptomatic illness rates among pregnant women and infants <6 months each season, and so we instead assumed these rates were equal to seasonal rates for adults 18–49 years and children 0–4 years, respectively^27^. We then calculated rates of symptomatic illness for each month by multiplying the seasonal rate by the proportion of FluSurv-NET hospitalizations that occurred that month, i.e., we assumed that symptomatic illnesses followed the same temporal distribution as hospitalizations. To estimate influenza-associated medically attended illnesses, we used previously published estimates of the fraction of symptomatic illnesses that are medically attended among adults 18–49 years and children 0–4 years and assumed that these were representative of the respective fractions among pregnant women and infants <6 months^28^.

Next, we obtained annual influenza vaccination coverage estimates for pregnant women aged ≥18 years from FluVaxView (Table S1)^13^. These estimates were generated from survey information on the percentage of women reporting influenza vaccination during the 12 months preceding a live birth. As monthly estimates were not available, we assumed that the monthly distribution of vaccines was equivalent to that of all adults 18–49 years at higher-risk of influenza-associated complications^29^. This group includes pregnant women and people with medical conditions associated with higher risk of influenza complications, such as asthma and heart disease. Infants <6 months are not eligible for vaccination but receive protection from maternal vaccination through the transplacental transfer of maternal antibodies. We assumed the probability that an infant was born to a vaccinated mother was equal to the cumulative vaccination coverage among pregnant women during the month of the infant’s birth.

In addition to vaccination coverage, we obtained estimates of influenza vaccine effectiveness (VE). We first collated season-specific estimates for women 18–49 years from the US Influenza Vaccine Effectiveness (Flu VE) Network (Figure S2)^30^. These estimates represent VE against medically attended outpatient illness and, given similar seroprotection rates among pregnant and non-pregnant women following influenza vaccination^31^, we assumed they were representative of influenza VE among pregnant women. We also assumed that infants <6 months exposed to maternal vaccination in the current season experienced the same VE as pregnant women, following evidence for efficient transplacental transfer of maternal influenza antibodies and comparable VE estimates between mothers and their infants^10,32–35^. As a sensitivity analysis, we instead assumed VE estimates for children 6 months–4 years obtained from the Flu VE Network were representative of VE among infants <6 months. These estimates were consistently higher than those used in the main analysis but were almost all within a 33–59% pooled confidence interval for VE among infants <6 months calculated in a prior meta-analysis (Figure S2)^11^. In all analyses, we conservatively set VE to 0% among infants whose mothers only received a prior season’s influenza vaccine, assuming a poor match between circulating strains and prior vaccine strains^36^. We also excluded possible protection gained postpartum through maternal influenza antibodies in breastmilk^37,38^.

Finally, we assumed that influenza virus infection during pregnancy conferred 100% protection to both the pregnant woman and her infant for the remainder of that season. Similar to vaccination, the probability that an infant was born to a previously infected mother was equal to the cumulative incidence of infection among pregnant women during the month of the infant’s birth.

### Framework to estimate influenza disease burden

We adapted a previously published multiplier model to estimate influenza disease burden among pregnant women and their infants <6 months each season (Figure 1A)^18,39,40^. First, we calculated monthly observed influenza-associated hospitalization rates for each group by dividing the monthly number of FluSurv-NET hospitalizations by the corresponding estimated catchment population and then adjusting for under-ascertainment using the under-detection multipliers. To derive the number of influenza-associated hospitalizations among all pregnant women and infants <6 months in the United States, we multiplied the corresponding adjusted hospitalization rates from FluSurv-NET by our US population estimates for each group. To estimate the total number of influenza-associated ICU admissions, we multiplied the estimated number of influenza-associated hospitalizations among each group by the corresponding proportion of FluSurv-NET hospitalizations that were admitted to the ICU that season. Finally, to estimate the total number of influenza-associated symptomatic illnesses, we multiplied our US population estimates by the estimated monthly rates of symptomatic illness. We then multiplied the number of symptomatic illnesses by the fraction that were medically attended to estimate the total number of influenza-associated medically attended illnesses. Our demographic and multiplier inputs are provided in Tables S1–S2.

**Figure 1.**
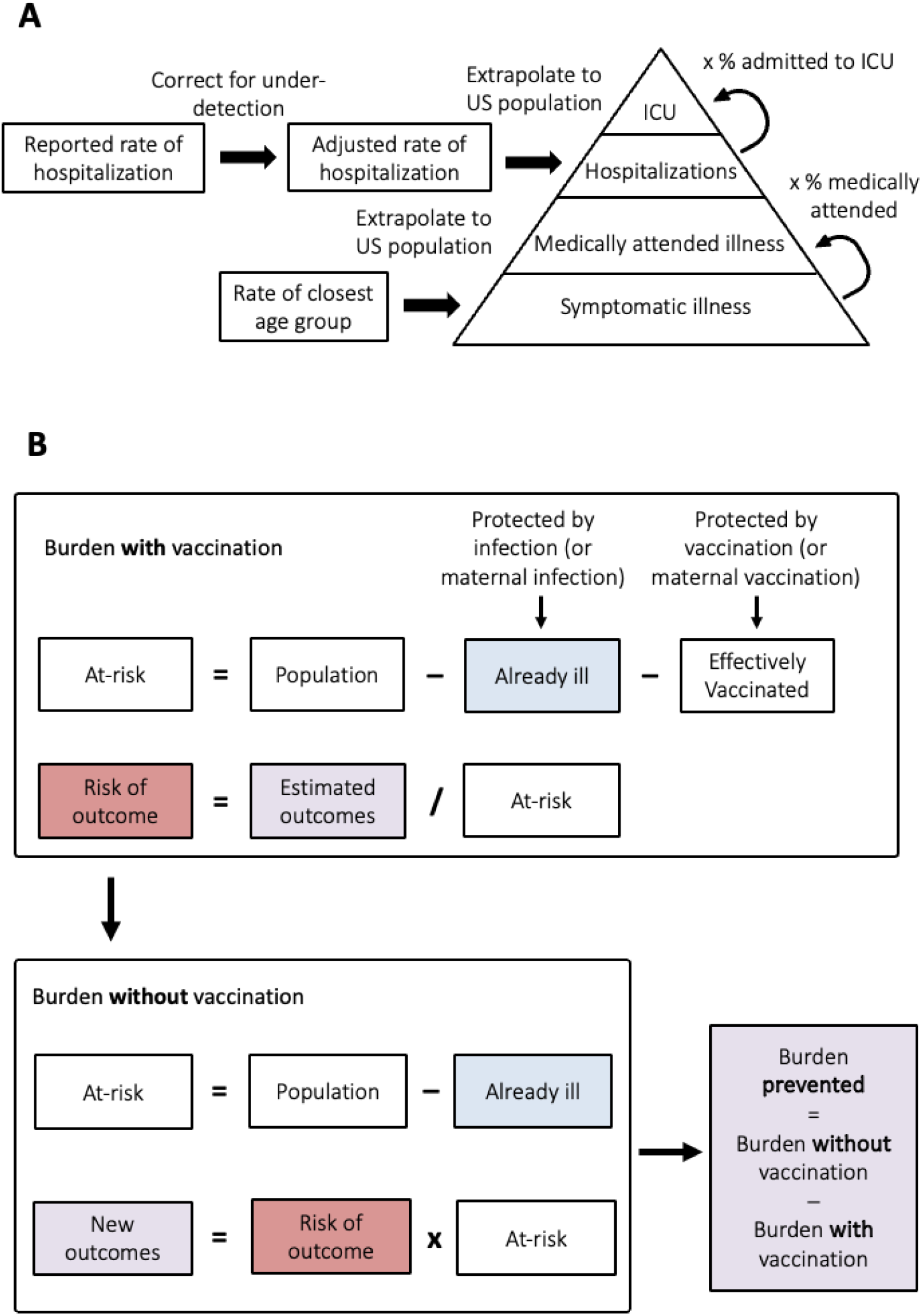
Frameworks for estimating influenza disease burden and burden prevented by influenza vaccination. (A) Multiplier model for estimating disease burden for each season and group (pregnant women or infants <6 months). (B) Compartmental framework for estimating disease burden prevented by vaccination for each season and group. The procedure is performed iteratively for each month of the season.

### Framework to estimate burden prevented by influenza vaccination

We adapted a previously published compartmental framework to estimate the burden prevented by maternal influenza vaccination among pregnant women and their infants <6 months each season (Figure 1B)^17,39,41,42^. This framework integrates vaccination coverage and effectiveness information with monthly symptomatic illness estimates (calculated above and henceforth referred to as ‘infections’) to track the number of people who were effectively vaccinated or infected, and thus protected from future illness, over the course of a season. For pregnant women, we calculated the monthly number who were effectively vaccinated as the population who had not yet been infected during the season multiplied by the monthly incident vaccination coverage and by VE (assuming vaccine-mediated protection is ‘all-or-nothing’). The remaining susceptible (or, ‘at-risk’) population was the number who had not yet been infected or effectively vaccinated. For infants <6 months, we applied the probability of being born to an infected mother (described above) to the monthly number of births to estimate the number protected by maternal infection. Similarly, we used the probability of being born to a vaccinated mother, multiplied by maternal VE, to estimate the number effectively protected by maternal vaccination. The susceptible population <6 months was then estimated as the number of infants born in the current and previous 5 months who were not protected by maternal infection or vaccination in the current season and who had not yet been infected.

For both pregnant women and infants <6 months, the respective susceptible populations were used to infer the monthly risk of each influenza disease burden outcome. For example, the monthly risk of symptomatic illness was calculated as the number of symptomatic illnesses estimated in the current month divided by the number susceptible in the previous month. Similarly, the monthly risks of medically attended illness, hospitalization, and ICU admission were calculated by dividing the corresponding number estimated in the current month by the previous month’s susceptible population. We then estimated the number of outcomes that would have occurred in the absence of vaccination by multiplying the monthly risk of each outcome by an adjusted susceptible population in which no one was protected by vaccination, i.e., no pregnant women were effectively vaccinated and no infants were protected by maternal vaccination. The number of influenza outcomes prevented by vaccination was then calculated as the difference between the number occurring in the presence of vaccination (i.e., the estimated disease burden outcomes) and the estimated number occurring without vaccination.

Uncertainty distributions for each influenza disease burden and vaccine-prevented burden outcome were generated using Monte Carlo simulation methods (Supplementary Information, Table S3)^43–45^. We also calculated the fraction of severe outcomes prevented as the number of influenza-associated hospitalizations prevented by vaccination divided by the total number that would have occurred without vaccination.

### Alternative influenza vaccination scenarios

We estimated the number of additional hospitalizations that could have been prevented under three alternative influenza vaccination scenarios that were designed to examine the potential impacts of increased and/or earlier vaccination uptake among pregnant women. In each season, the first scenario assumed the timing of vaccination remained the same, but that final coverage reached the Healthy People 2030 objective of 70%; the second scenario assumed that timing was shifted one month earlier (with July being the earliest start month) but that final coverage did not change; and the third scenario combined the first and second scenarios, i.e., timing was shifted one month earlier and final coverage reached 70% (Figure S3).

All analyses were conducted in R 4.2.0^46^, and final estimates are reported to three significant figures unless specified otherwise. This activity was reviewed by CDC, deemed not research, and was conducted consistent with applicable federal law and CDC policy: 45 C.F.R. part 46.102(l)(2), 21 C.F.R. part 56; 42 U.S.C. Sect. 241(d); 5 U.S.C. Sect. 552a; 44 U.S.C. Sect. 3501 et seq. All data were the result of secondary analyses and used solely as model inputs. FluSurv-NET sites obtained human subjects and ethics approval from their respective state health department, academic partner, and participating hospital institutional review boards.

## Results

From 2011/12–2019/20, there were approximately 3 million pregnant women and 2 million infants <6 months in the United States in any given season (Tables S1–S2). Across individual seasons, we estimated there were between 83,000–310,000 influenza-associated symptomatic illnesses, 30,700–115,000 medically attended illnesses, 2,670–10,000 hospitalizations, and 76–486 ICU admissions among pregnant women (Figure 2, Figure S4). Rates of hospitalization per 100,000 were 1.56–6.81 times higher among pregnant women than corresponding estimates for all adults 18–49 years that were generated using similar methodology (Table S4)^47^. Infants <6 months generally experienced greater total disease burden than pregnant women, with 91,800–361,000 symptomatic illnesses, 61,500–242,000 medically attended illnesses, 3,960–13,100 hospitalizations, and 457–1,830 ICU admissions (Figure 2, Figure S4). Rates of hospitalization per 100,000 were 2.79–6.16 times higher than corresponding estimates for children 0–4 years^47^, and higher than corresponding estimates for pregnant women (Table S4).

**Figure 2.**
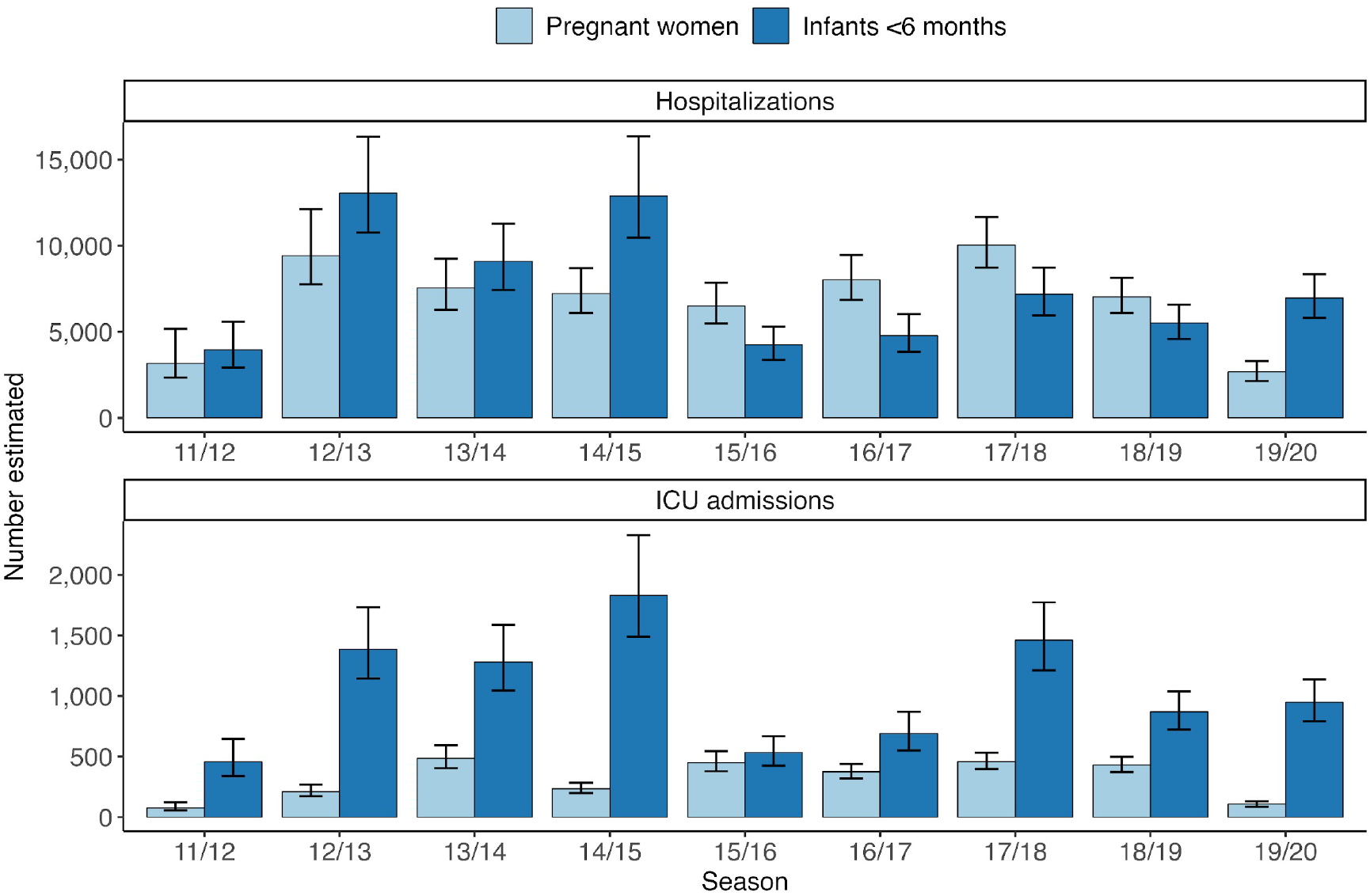
Estimated influenza-associated hospitalizations and ICU admissions among pregnant women and infants <6 months from 2011/12–2019/20. Bars show point estimates and error bars are the 95^th^ percentile uncertainty intervals.

Influenza vaccination coverage among pregnant women ranged from 47% in 2011/12 to 61% in 2019/20, and VE against medically attended influenza in women 18–49 years ranged from 1% in 2014/15 to 54% in 2013/14 (Table S1, Figure S2). Across individual seasons, we estimated that influenza vaccination prevented between 933–99,600 influenza-associated symptomatic illnesses, 345–36,800 medically attended illnesses, 33–2,440 hospitalizations, and 1–166 ICU admissions among pregnant women (Figure 3, Figure S5). The number of prevented hospitalizations was equivalent to 0.45–27% of influenza-associated hospitalizations that would have occurred in the absence of vaccination (Figure S6).

**Figure 3.**
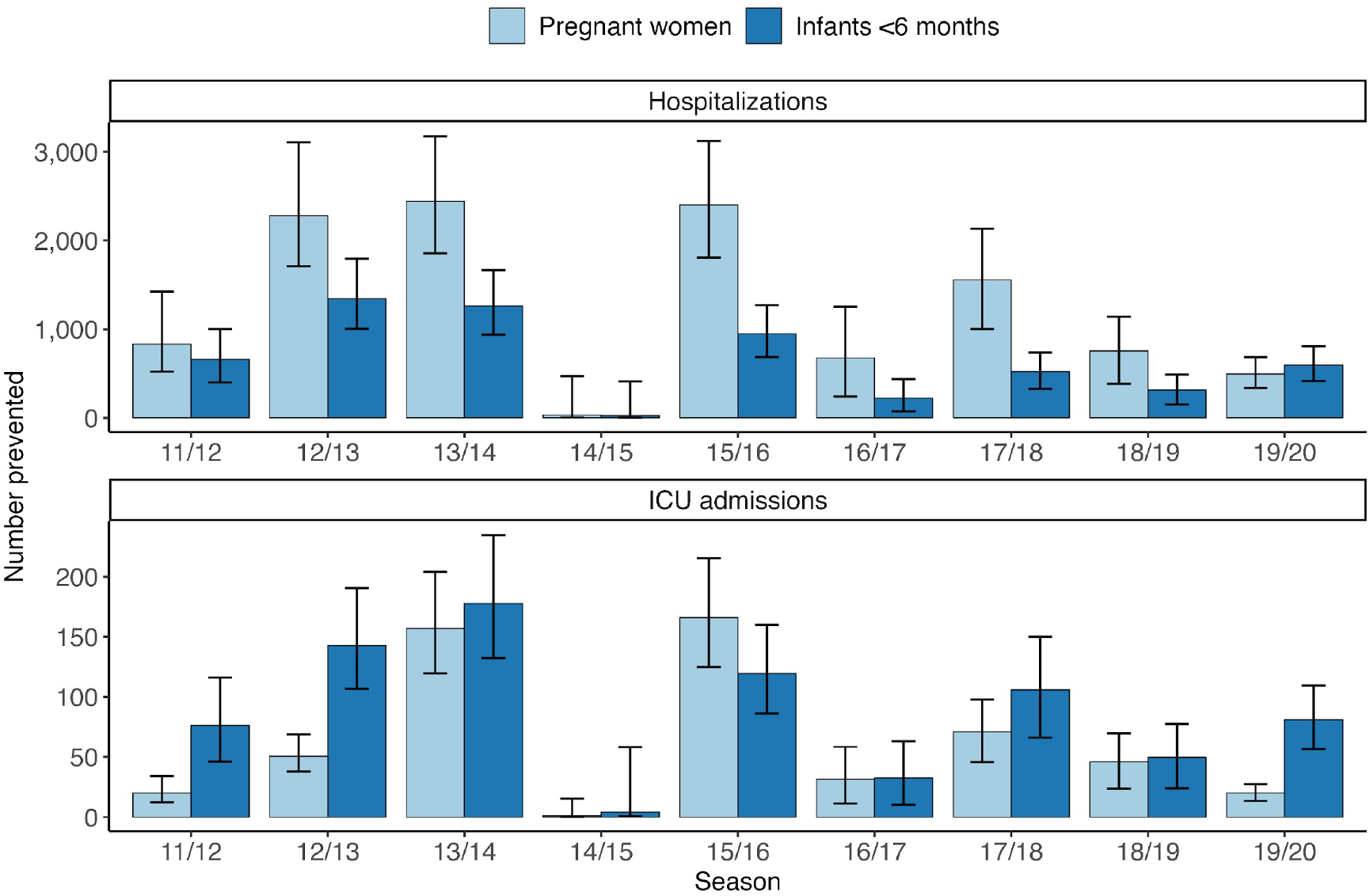
Estimated influenza-associated hospitalizations and ICU admissions prevented by maternal influenza vaccination among pregnant women and infants <6 months from 2011/12–2019/20. Bars show point estimates and error bars are the 95^th^ percentile uncertainty intervals.

Among infants <6 months, maternal influenza vaccination prevented an estimated 733–48,800 symptomatic illnesses, 491–32,700 medically attended illnesses, 29–1,350 hospitalizations, and 4–178 ICU admissions (Figure 3, Figure S5). These estimates were generally 1–3 times higher when assuming infants <6 months experienced the same VE as children 6 months–4 years, except in 2013/14 when estimates were 40 times higher due to VE of just 1% among women 18–49 years (Figures S2 and S7). Although vaccination typically prevented more hospitalizations among pregnant women, more ICU admissions were generally prevented among infants <6 months. This was due to the higher proportion of hospitalizations admitted to the ICU among infants <6 months compared to pregnant women (Tables S1–S2). Overall, 0.22–18.3% of influenza-associated hospitalizations were prevented among infants <6 months (Figure S6).

With respect to alternative influenza vaccination scenarios, we estimated that greater vaccination uptake would have prevented more influenza-associated hospitalizations among pregnant women than earlier vaccination uptake, with the latter generally having little or no impact on prevented outcomes (Figure 4). In contrast, earlier vaccination uptake typically prevented more hospitalizations among infants <6 months than greater vaccination uptake. One exception was 2011/12, when there was very late influenza transmission (peaking in March) and low vaccination coverage among pregnant women (Table S1)^48^. Compared to observed vaccination coverage, greater and earlier uptake could have prevented an additional 12–942 hospitalizations among pregnant women and 17–1,010 among infants <6 months across individual seasons. This corresponded to an absolute total of 45–3,350 prevented hospitalizations among pregnant women and 46–2,350 among infants <6 months across seasons (Figure S8).

**Figure 4.**
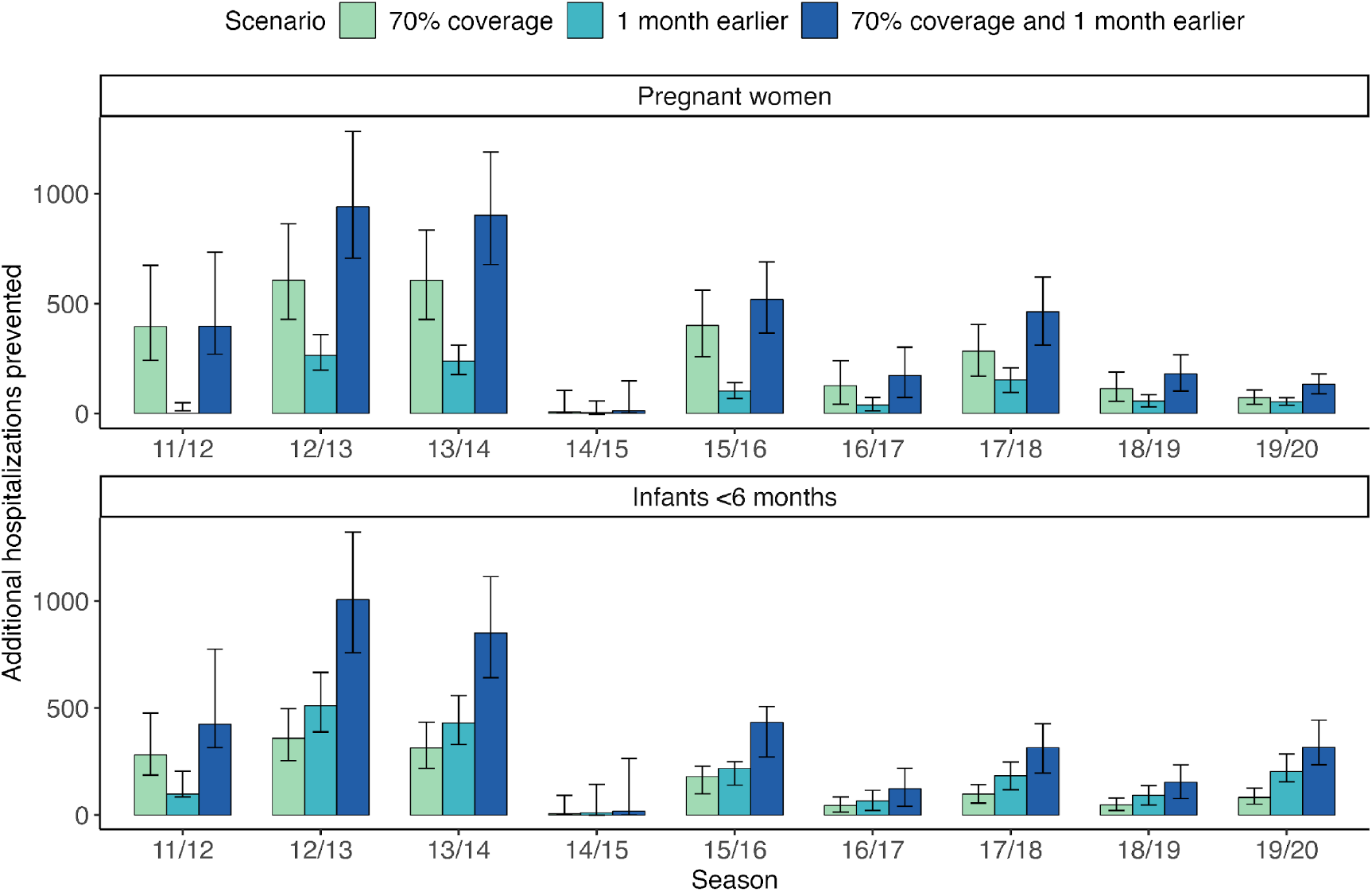
Additional influenza-associated hospitalizations prevented among pregnant women and infants <6 months if maternal influenza vaccination coverage was increased to 70% and/or was shifted one month earlier. In the scenarios including earlier coverage, vaccination was started no earlier than July. Bars show point estimates and error bars are the 95^th^ percentile uncertainty intervals.

## Discussion

Pregnant women and infants <6 months are at increased risk of influenza-associated complications. Influenza vaccination during pregnancy has a strong safety record and is effective in reducing the risk of illness in these groups^10,11,49–53^. We extended a previously published framework to estimate influenza disease burden, and the burden prevented by maternal influenza vaccination, among pregnant women and their infants <6 months from 2011/12–2019/20. We estimated substantial disease burden among both groups over the study period, with higher influenza-associated hospitalization rates than the closest age group in the general population with corresponding estimates (all adults 18–49 years and children 0–4 years, respectively)^47^. Our estimates also indicated that influenza vaccination was effective in preventing influenza-associated hospitalizations and ICU admissions among both pregnant women and their infants <6 months. This aligns with previous findings that maternal influenza vaccination is cost-effective for these groups^54–56^ and supports its important public health benefits.

While influenza vaccination coverage among pregnant women ≥18 years increased during the study period (and was higher than coverage among all adults 18–49 years at higher risk of influenza complications), it has dropped substantially following the onset of the COVID-19 pandemic in 2020, with one estimate among pregnant women 18–49 years as low as 38% during the 2024/25 influenza season^13,57^. Our analysis of additional vaccination scenarios suggested that increases in influenza vaccination uptake could have prevented additional disease burden among pregnant women and their infants <6 months. However, this also implies that significant declines in uptake, such as those occurring since 2020, could lead to substantial increases in influenza disease burden among both groups. Therefore, encouraging uptake of maternal influenza vaccination is crucial to reducing illness in pregnant women and infants <6 months. Given that the odds of influenza vaccination increase when pregnant women believe it will benefit themselves or their baby^58^, providers should include discussion of the benefits of vaccination to both mother and baby when counseling pregnant women about influenza vaccination.

In addition to influenza vaccination coverage levels, the timing of vaccination impacts the protection experienced by pregnant women and their infants^15^. Previous work has shown that the timing of vaccination in the United States is primarily aligned to protect pregnant women and may occur too late for infants born before the beginning of the vaccination campaign (e.g., infants born in the summer months) whose first 6 months of life will overlap with the upcoming influenza season^16^. This is consistent with our finding that earlier influenza vaccination uptake each season could have prevented more hospitalizations among infants <6 months than higher uptake, whereas the converse was true for pregnant women. These results support the consideration of vaccination in July or August for pregnant women in their third trimester to reduce the risk of illness in their infants after birth. Although early vaccination for pregnant women in their first or second trimester may not be advised due to the risk of waning vaccine protection over the course of the influenza season, there are alternative strategies to help protect infants <6 months born before the vaccination campaign, including influenza vaccination of household contacts and caregivers^7,59^.

Although a compartmental framework has been used previously to estimate the population-level benefits of influenza vaccination for pregnant women^60^, we show how such analyses can be extended to estimate corresponding benefits for their infants <6 months. Our framework provides a means of evaluating the benefits of influenza vaccination among two groups at increased risk of influenza-associated complications and may be useful for other countries considering the expansion or evaluation of existing maternal influenza vaccination programs. Once data from more recent seasons become available, our work could also be extended to describe the impact of maternal influenza vaccination following declines in coverage since 2020, and the impact of other seasonal respiratory vaccines administered during pregnancy, such as vaccines against respiratory syncytial virus.

Our approach is subject to some limitations. First, FluSurv-NET covers approximately 9% of the US population and may not be generalizable to the entire country. Second, we assumed equal annual mortality rates among infants <6 months and >6 months, and our population estimates will be overestimated if mortality rates are higher among the former^61^. Although this may impact our absolute estimates of influenza disease burden and burden prevented by vaccination, our estimates of the rates of disease burden per 100,000 and the percentage of burden prevented by vaccination should be more robust to changes in the underlying population. Similarly, the multipliers for under-reporting, rates of symptomatic illness, and fraction of cases that are medically attended were not specific to pregnant women or infants <6 months, and instead reflected the closest populations for which data were available (typically, adults 18–49 years and children 0–4 years, respectively). If pregnant women and infants <6 months are more likely to be tested for influenza in hospital settings, we may have over-estimated the number of hospitalizations and ICU admissions. Conversely, we may have under-estimated the number of symptomatic and medically attended illnesses if these groups are more likely to experience influenza illness and receive medical attention compared with the general population. Determining the likelihood of medically attended illness is particularly challenging for pregnant women as routine antenatal appointments may influence whether and when they receive care for influenza-associated illness.

With respect to influenza vaccination, we assumed that the timing of vaccination among pregnant women was equivalent to that of all adults 18–49 years at higher risk of influenza complications. Our analysis of alternative vaccination scenarios suggested that vaccination earlier in the season may increase our estimates of prevented burden for infants <6 months but would have minimal impact on our estimates for pregnant women. Second, our final vaccination coverage data were based on vaccinations received in the 12 months preceding a woman’s recent live birth and may therefore include vaccinations received before pregnancy. This may lead to over-estimation of the proportion of infants <6 months protected by vaccination if vaccination must occur during pregnancy for the transplacental transfer of maternal antibodies. Third, we assumed the same VE among all infants <6 months. Although protection may decrease over the first 6 months of life^10,62–64^, our assumption that infant VE was equivalent to VE among pregnant women was based on estimates for infants <6 months and so should account for underlying heterogeneity by age. Further information would be needed to parameterize VE inputs by age and by season. Fourth, our VE estimates were based on VE against medically attended illness, which may be lower than VE against hospitalization and/or ICU admission and lead to under-estimation of vaccine-prevented hospitalizations and ICU admissions^62^. We also conservatively assumed that influenza infection during pregnancy conferred complete protection to the pregnant woman and her infant for the remainder of the season. Incomplete protection would lead to a greater at-risk population and therefore a greater estimated impact of vaccination. Finally, we employed a static model that assumes ‘all-or-nothing’ vaccine-mediated protection and does not incorporate indirect effects of vaccination through herd immunity. Dynamic models could be explored to address these issues but would require additional data on influenza transmission and natural history in pregnant women and young infants to inform model inputs.

In this work, we estimate the burden of influenza, and the burden prevented by maternal influenza vaccination, among pregnant women and their infants <6 months in the United States from 2011–2020. Our analyses suggest that influenza vaccination reduces disease burden among both groups and support its important public health benefits.

## Data Availability

All data were the result of secondary analyses and used solely as model inputs. Data available online are cited within the text.

## Acknowledgements

The authors would like to thank Kendall Williams and Amelia Heckmann for their contributions to exploratory data analysis.

## Disclaimer

The findings and conclusions in this report are those of the authors and do not necessarily represent the views of the Centers for Disease Control and Prevention.

