## Supplementary Information for "Public health benefits of maternal influenza vaccination among pregnant women and infants <6 months in the United States, 2011–2020"

#### Demographic information

Monthly numbers of live births were obtained from the National Vital Statistics System and annual mortality rates for infants <1 year per 1,000 live births were obtained from the National Center for Health Statistics (NCHS)[1,2]. Annual pregnancy rates per 1,000 females 15–44 years were also acquired from NCHS, in addition to the percentage of pregnancies ending in live birth, abortion, or fetal loss. Annual estimates of the total US population of females 15–44 years were obtained from the US Census Bureau[3].

#### Monte Carlo simulations

Uncertainty distributions for each influenza disease burden and vaccine-prevented burden outcome were generated using Monte Carlo simulation methods. For key input variables, we drew 10,000 samples from pre-defined probability distributions (Table S3) and re-estimated the number of disease burden outcomes and vaccine-prevented outcomes for each new sample. Uncertainty intervals were calculated as the 95th percentiles of the resulting distributions.

### Supplementary tables

| Season | Population | Hospitalization multiplier | ICU (%) | MA fraction | Symptomatic illness /100,000 | Vaccination coverage (%) |
|---|---|---|---|---|---|---|
| 11/12 | 3,238,639 | 3.54 (2.34-7.28) | 2.38 | 0.37 (0.35-0.40) | 2,564 (2,021-3,850) | 47.4 (46.6-48.2) |
| 12/13 | 3,232,779 | 2.83 (2.21-3.94) | 2.22 | 0.37 (0.35-0.40) | 8,383 (7,127-10,509) | 55.3 (54.5-56.1) |
| 13/14 | 3,210,481 | 2.46 (2.03-3.10) | 6.43 | 0.37 (0.35-0.40) | 9,589 (8,429-11,180) | 56.1 (55.3-56.9) |
| 14/15 | 3,258,009 | 1.96 (1.68-2.36) | 3.26 | 0.37 (0.35-0.40) | 6,310 (5,683-7,108) | 57.3 (56.5-58.2) |
| 15/16 | 3,241,606 | 2.44 (2.09-2.92) | 6.91 | 0.37 (0.35-0.40) | 6,667 (4,770-10,685) | 60.0 (59.2-60.9) |
| 16/17 | 3,218,341 | 2.30 (2.01-2.69) | 4.66 | 0.37 (0.35-0.40) | 6,786 (4,858-11,261) | 59.0 (58.2-59.8) |
| 17/18 | 3,142,778 | 1.80 (1.58-2.09) | 4.57 | 0.37 (0.35-0.40) | 9,931 (7,135-16,647) | 59.2 (58.5-59.9) |
| 18/19 | 3,093,181 | 1.69 (1.51-1.92) | 6.11 | 0.37 (0.35-0.40) | 7,088 (5,214-12,685) | 60.8 (60.0-61.6) |
| 19/20 | 3,058,022 | 1.68 (1.45-2.00) | 4.00 | 0.37 (0.35-0.40) | 10,423 (6,816-29,185) | 61.1 (60.3-61.9) |

**Table S1 – Input parameters for pregnant women.** Parentheses denote 95% confidence intervals for parameters included in the Monte Carlo sampling procedure. Abbreviations: ICU = intensive care unit; MA = medically attended.

| Season | Population | Hospitalization multiplier | ICU (%) | MA fraction | Symptomatic illness /100,000 |
|---|---|---|---|---|---|
| 11/12 | 1,968,957 | 2.34 (1.80-3.33) | 11.54 | 0.67 (0.64-0.70) | 4,697 (3,750-6,248) |
| 12/13 | 1,969,112 | 1.86 (1.53-2.36) | 10.61 | 0.67 (0.64-0.70) | 17,835 (11,570-41,651) |
| 13/14 | 1,965,918 | 1.90 (1.60-2.33) | 14.08 | 0.67 (0.64-0.70) | 12,712 (8,894-22,319) |
| 14/15 | 1,988,996 | 2.17 (1.76-2.83) | 14.23 | 0.67 (0.64-0.70) | 16,136 (10,813-31,803) |
| 15/16 | 1,980,868 | 1.58 (1.38-1.85) | 12.61 | 0.67 (0.64-0.70) | 11,028 (7,619-21,415) |
| 16/17 | 1,955,397 | 1.70 (1.46-2.04) | 14.40 | 0.67 (0.64-0.70) | 11,950 (6,966-62,524) |
| 17/18 | 1,919,629 | 1.63 (1.43-1.89) | 20.30 | 0.67 (0.64-0.70) | 16,867 (12,158-30,462) |
| 18/19 | 1,884,162 | 1.45 (1.31-1.62) | 15.79 | 0.67 (0.64-0.70) | 14,714 (11,939-20,602) |
| 19/20 | 1,863,539 | 1.45 (1.28-1.66) | 13.62 | 0.67 (0.64-0.70) | 17,796 (14,165-32,489) |

**Table S2 – Input parameters for infants <6 months.** Parentheses denote 95% confidence intervals for parameters included in the Monte Carlo sampling procedure. Population size represents the mean population of infants <6 months across all months in the season. Abbreviations: ICU = intensive care unit; MA = medically attended.

| Input | Stratifications | Distribution | Parameter(s) |
|---|---|---|---|
| FluSurv-NET hospitalizations | Season, month, group | Poisson | $\lambda$ = observed number |
| Hospitalization multiplier | Season, group | Beta-PERT | $m$ = observed value<br>$m_l$ = observed lower 95th percentile<br>$m_l$ = observed upper 95th percentile |
| Symptomatic illness rate | Season, month, group | Normal | $\mu$ = observed rate<br>$\sigma$ = observed SE |
| Medically attended fraction | Season, group | Normal | $\mu$ = observed fraction<br>$\sigma$ = observed SE |
| Vaccination coverage | Season, month | Normal | $\mu$ = observed coverage<br>$\sigma$ = observed SE |
| Logarithm of 1−VE | Season, month, group | Normal | $\mu$ = observed log(1−VE)<br>$\sigma$ = observed SE of log(1−VE) |

**Table S3 – Monte Carlo simulation settings.** Parameter notation is as follows: $\lambda$ reflects the mean of a Poisson distribution; $N$ and $p$ reflect the number of trials and probability of success of a Binomial distribution; $\mu$ and $\sigma$ reflect the mean and standard deviation of a Normal distribution; and $m$, $m_l$ and $m_u$ reflect the mode, minimum and maximum of a Beta-PERT distribution. Normal distributions were truncated at 0. Stratification by 'group' refers to pregnant women or infants <6 months. Abbreviations: SE = standard error; VE = vaccine effectiveness.

| Season | Pregnant women | Adults 18-49 years | Ratio (pregnant women/ adults 18-49 years) | Infants <6 months | Children 0-4 years | Ratio (Infants <6 months/ children 0-4 years) |
|---|---|---|---|---|---|---|
| 11/12 | 98.02 | 14.40 | 6.81 | 201.28 | 32.70 | 6.16 |
| 12/13 | 291.61 | 47.10 | 6.19 | 663.26 | 124.20 | 5.34 |
| 13/14 | 235.30 | 53.80 | 4.37 | 462.75 | 88.60 | 5.22 |
| 14/15 | 221.57 | 35.40 | 6.26 | 647.50 | 112.50 | 5.76 |
| 15/16 | 200.70 | 37.40 | 5.37 | 214.18 | 76.90 | 2.79 |
| 16/17 | 249.12 | 38.10 | 6.54 | 245.07 | 83.30 | 2.94 |
| 17/18 | 319.29 | 55.30 | 5.77 | 374.67 | 117.60 | 3.19 |
| 18/19 | 227.29 | 41.20 | 5.53 | 292.12 | 102.60 | 2.85 |
| 19/20 | 87.30 | 55.90 | 1.56 | 373.97 | 124.10 | 3.01 |

**Table S4** – Hospitalization rates per 100,000 among pregnant women and infants <6 months compared to estimates among the closest age group in the general population (source cited in the main text). Values are reported to two decimal places.

### Supplementary figures

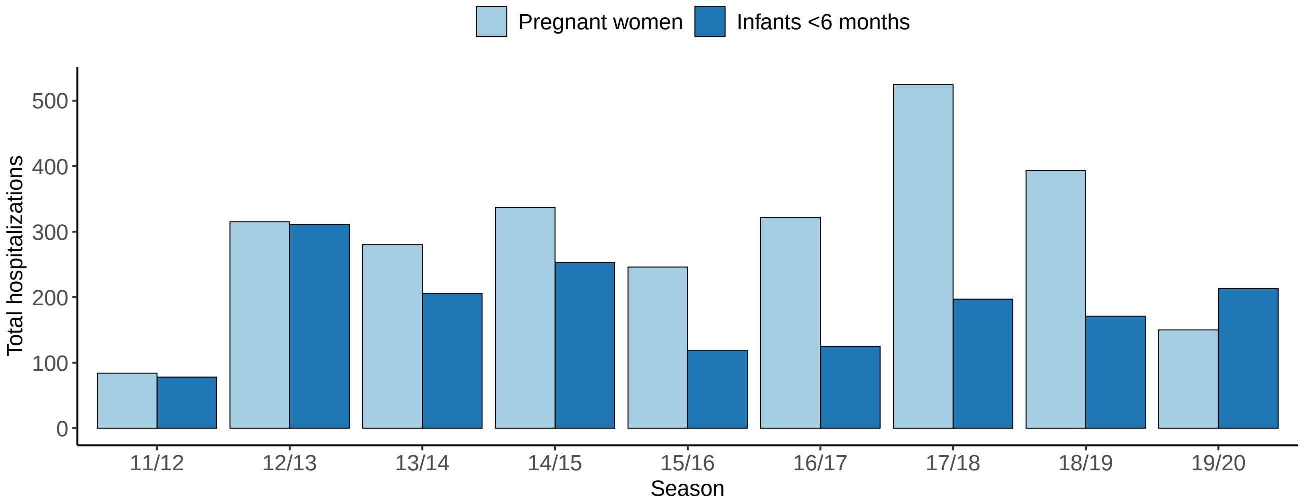


**Figure S1 – Total influenza-associated hospitalizations reported each season in FluSurv-NET.** Bars indicate reported data, before adjusting for under-ascertainment and catchment population size.

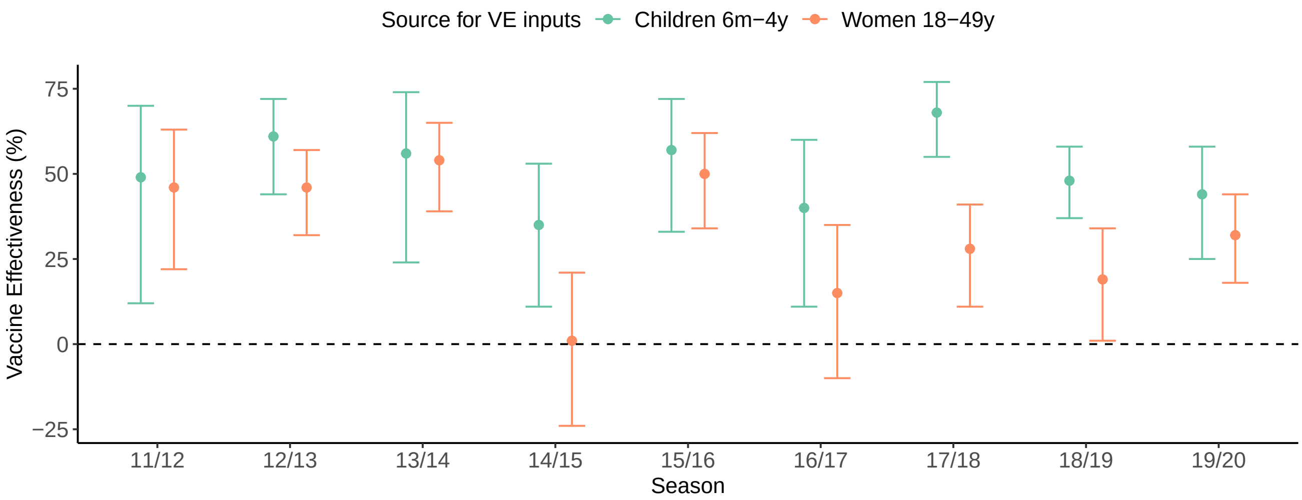


**Figure S2 – Influenza VE inputs.** In the main analysis, estimates among all women 18–49 years were used to parameterize VE for pregnant women and infants <6 months. In sensitivity analyses, we instead used estimates among children 6 months–4 years to parameterize VE for infants <6 months. Error bars indicate 95% confidence intervals around the point estimates. Abbreviations: VE = vaccine effectiveness.

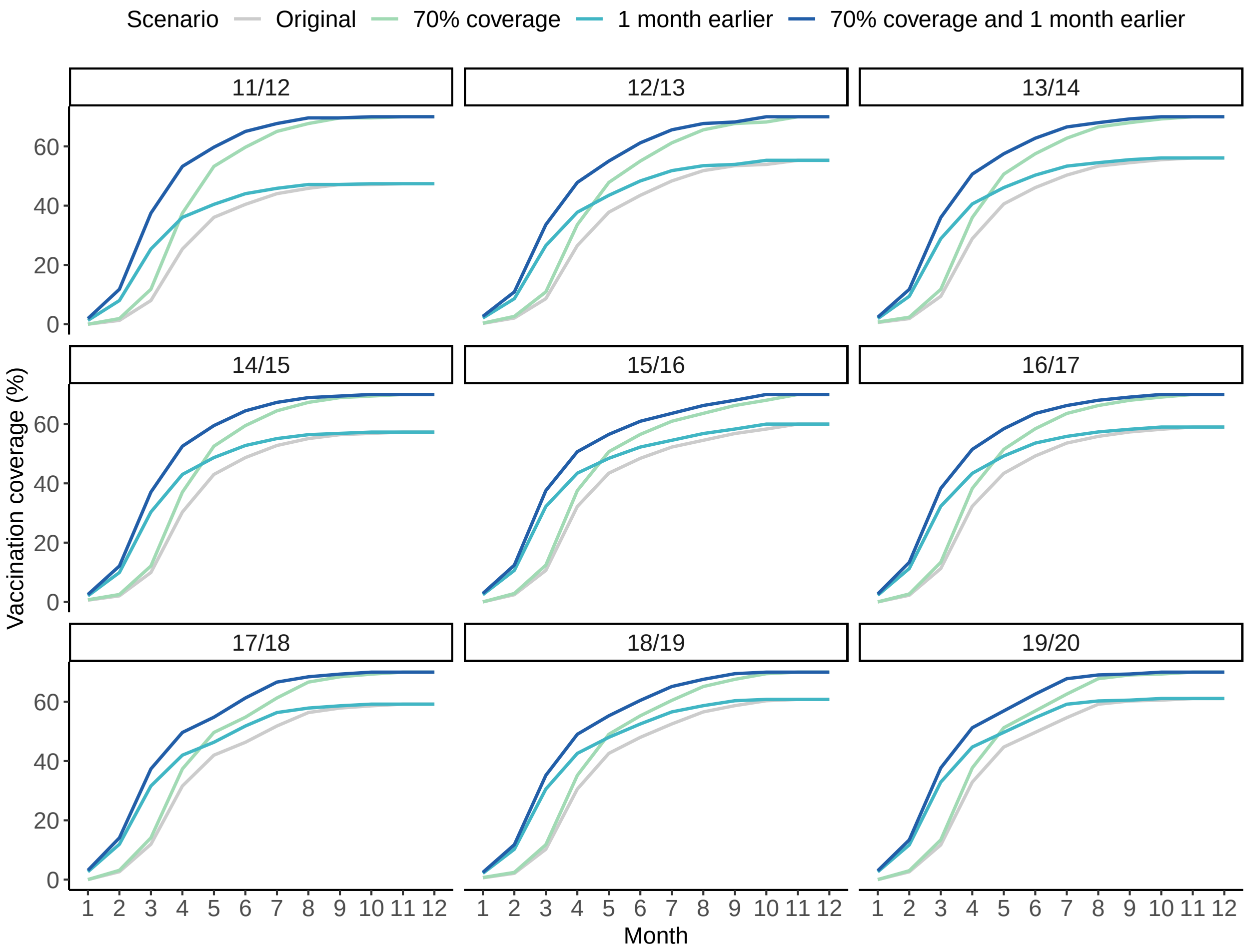


**Figure S3 – Original and alternative influenza vaccination coverage scenarios for the 2011/12 to 2019/20 influenza seasons.** The original scenario represents observed vaccination coverage among pregnant women. Alternative scenarios represent coverage that is increased to 70%, coverage that is shifted 1 month earlier, and coverage that reaches 70% and is shifted 1 month earlier. Months are relative to the beginning of the original vaccination campaign (July is 1, August is 2, and so on).

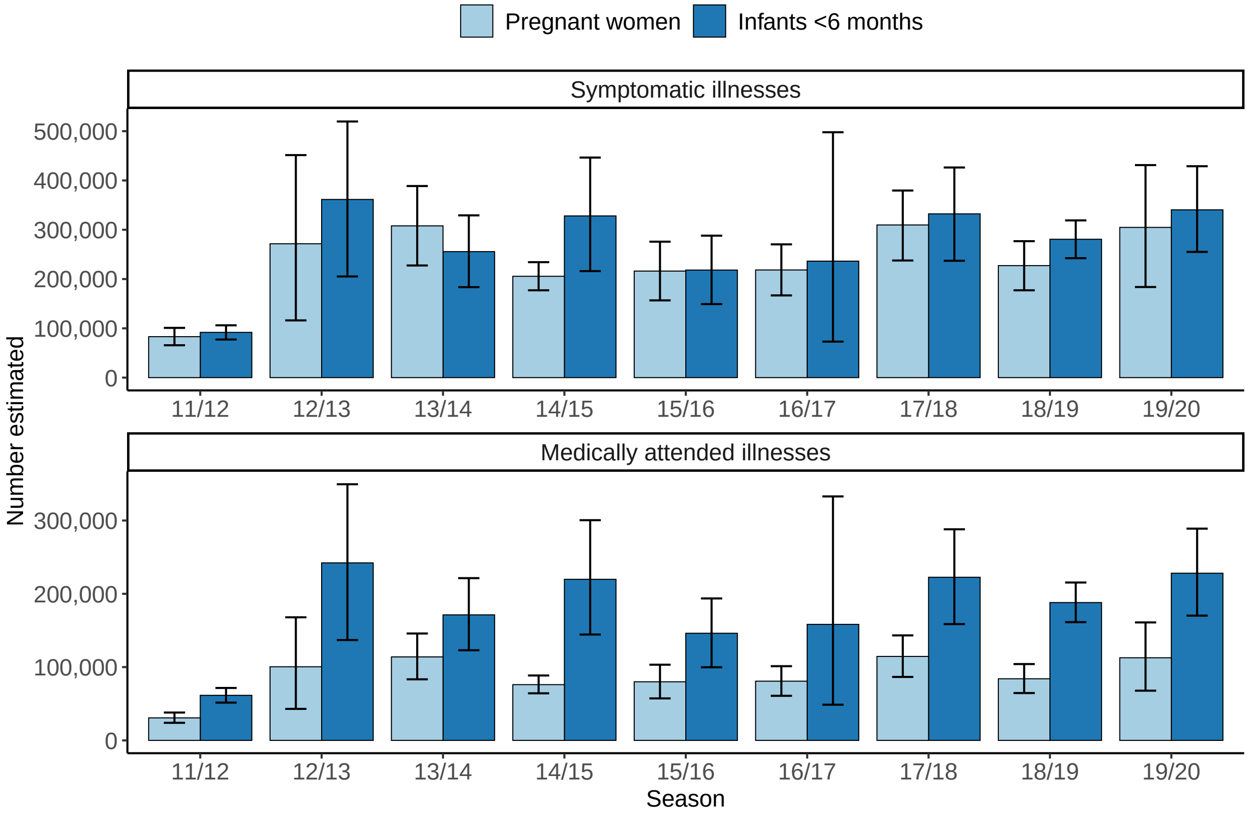


**Figure S4 – Estimated influenza-associated symptomatic and medically attended illnesses among pregnant women and infants <6 months from 2011/12–2019/20.** Bars show point estimates and error bars are the 95th percentile uncertainty intervals.

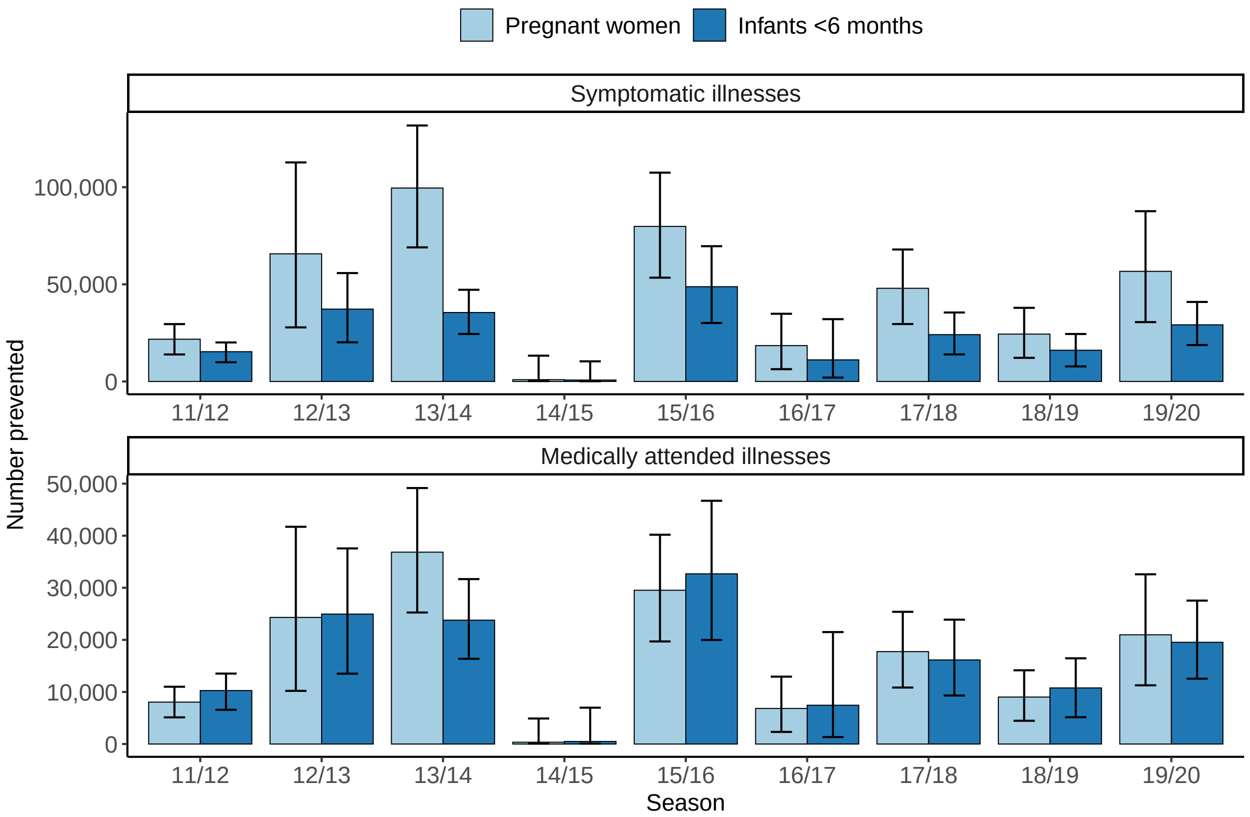


**Figure S5 – Estimated influenza-associated symptomatic and medically attended illnesses prevented by maternal influenza vaccination among pregnant women and infants <6 months from 2011/12–2019/20.** Bars show point estimates and error bars are the 95th percentile uncertainty intervals.

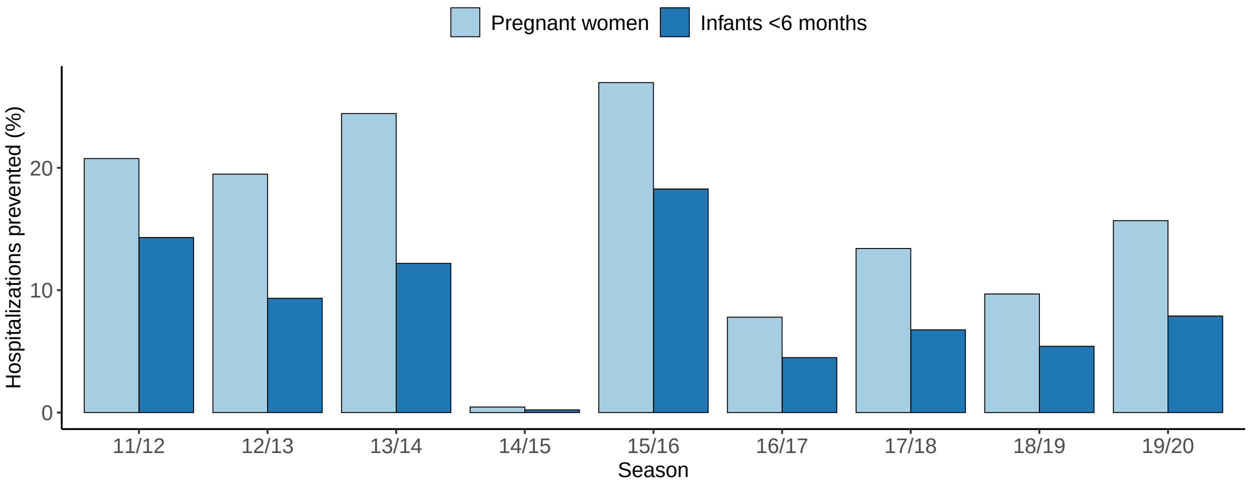


**Figure S6 – Estimated percentage of influenza-associated hospitalizations prevented by maternal influenza vaccination among pregnant women and infants <6 months from 2011/12–2019/20.** Bars show point estimates.

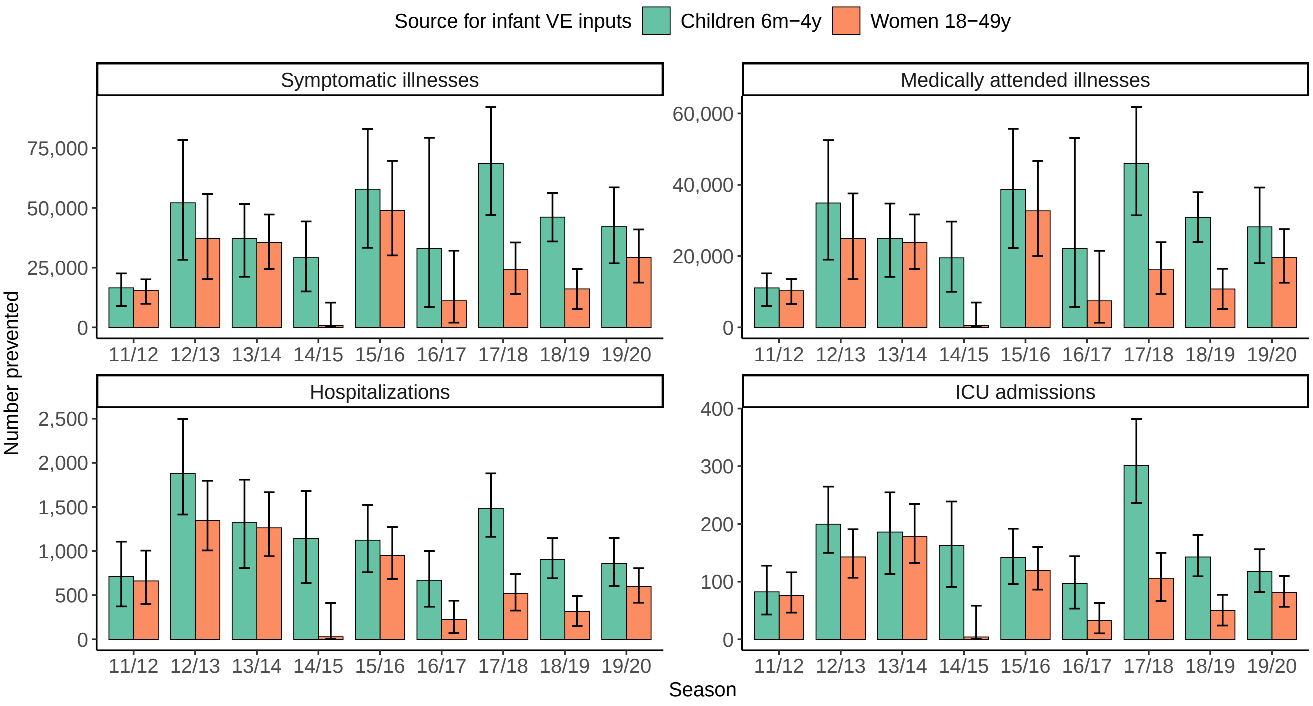


**Figure S7 – Estimated number of influenza-associated outcomes prevented by maternal influenza vaccination among infants <6 months under different infant VE assumptions.** VE estimates among all women 18–49 years were used for the main analysis and estimates among children 6 months–4 years were used as a sensitivity analysis. Bars show point estimates and error bars are the 95th percentile uncertainty intervals. Abbreviations: VE = vaccine effectiveness.

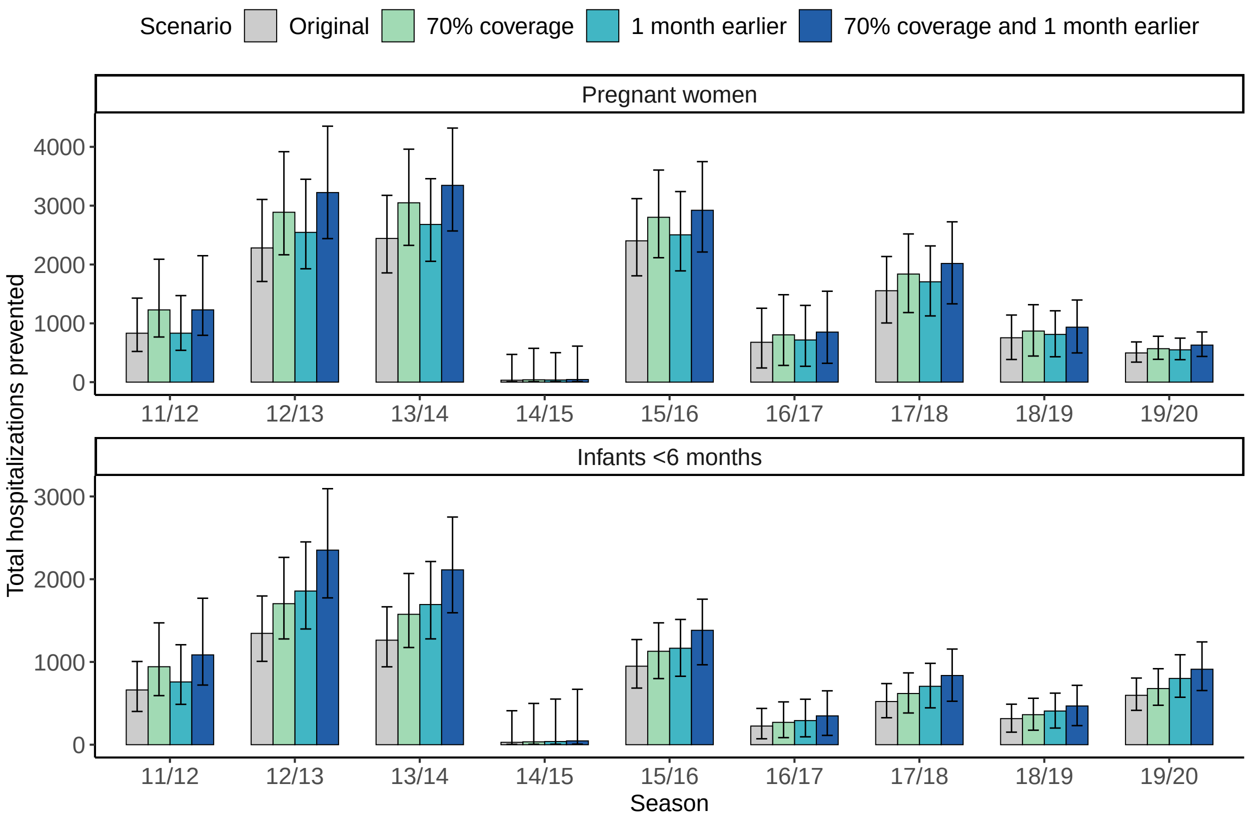


**Figure S8 – Total influenza-associated hospitalizations prevented among pregnant women and infants <6 months if influenza vaccination coverage among pregnant women was increased to 70% and/or shifted one month earlier.** The original estimates with observed vaccination coverage are shown in grey for comparison. Bars show point estimates and error bars are the 95th percentile uncertainty intervals.
